# Impact of Mydriasis on Image Gradability and Automated Diabetic Retinopathy Screening with a Handheld Camera in Real-World Settings

**DOI:** 10.1101/2025.01.02.25319898

**Authors:** Iago Diogenes, David Restrepo, Lucas Zago Ribeiro, Andre Kenzo Aragaki, Fernando Korn Malerbi, Caio Saito Regatieri, Luis Filipe Nakayama

**Affiliations:** Sao Paulo Federal University, Ophthalmology Department, Sao Paulo – SP, Brazil; Laboratory of Mathematics and Computer Science (MICS), Centrale Supélec, Paris-Saclay University, Gif-sur-Yvette, France; Laboratory for Computational Physiology, Massachusetts Institute of Technology, Cambridge, MA, USA

## Abstract

**Purpose:** Diabetic retinopathy (DR) screening in low- and middle-income countries (LMICs) faces challenges due to limited access to specialized care. Portable retinal cameras provide a practical alternative, but image quality, influenced by mydriasis, affects artificial intelligence (AI) model performance. This study examines the role of mydriasis in improving image quality and AI-based DR detection in resource-limited settings.

**Methods:** We compared the proportion of gradable images between mydriatic and non-mydriatic groups and used logistic regression to identify factors influencing image gradability, including age, gender, race, diabetes duration, and systemic hypertension. A ResNet-200d algorithm was trained on the mBRSET dataset and validated on mydriatic and non-mydriatic images. Performance metrics, such as accuracy, F1 score, and AUC, were evaluated.

**Results:** The mydriatic group had a higher proportion of gradable images (82.1% vs. 55.6%, *P*< 0.001). Factors such as systemic hypertension, older age, male gender, and longer diabetes duration were associated with lower image gradability in non-mydriatic images. Mydriatic images achieved better AI performance, with accuracy (82.91% vs. 79.23%), F1 score (0.83 vs. 0.79), and AUC (0.94 vs. 0.93). Among gradable images, the performance difference was not statistically significant.

**Conclusion:** Mydriasis improves image gradability and enhances AI model performance in DR screening. However, optimizing AI for non-mydriatic imaging is critical for LMICs where mydriatic agents may be unavailable. Refining AI models for consistent performance across imaging conditions is essential to support the broader implementation of AI-driven DR screening in resource-constrained settings.

## Introduction

The global increase in the prevalence of retinal diseases, particularly diabetic retinopathy (DR), has highlighted the urgent need for effective screening and diagnostic tools, especially in low- and middle-income countries (LMICs) (Magliano & Boyko 2021, Yim et al. 2021). In these regions, access to specialized ophthalmological care is often limited, leading to delayed diagnoses, disease progression, and poorer patient outcomes (Yim et al. 2021). Portable retinal fundus cameras have emerged as a vital solution, offering a cost-effective means to improve access to retinal imaging, particularly in remote or underserved areas (de Oliveira et al. 2023, Rajalakshmi et al. 2021, Vujosevic et al. 2020). These devices provide a practical alternative to traditional tabletop cameras, enabling healthcare providers to capture retinal images in non-clinical settings (de Oliveira et al. 2023).

A key factor in obtaining high-quality retinal images is whether the patient’s pupils are dilated (mydriasis) or not (non-mydriatic) (Piyasena et al. 2018). Mydriasis, achieved with pharmacological agents, allows for a wider and clearer view of the retina, making it easier to detect subtle changes crucial for early diagnosis and management of diseases like DR (Piyasena et al. 2018). Non-mydriatic imaging, while more convenient as it avoids the need for eye drops and reduces patient discomfort, may compromise image quality, particularly in darker eyes or in sub-optimal lighting conditions (Banaee et al. 2017). Moreover, administering mydriatic eyedrops requires trained professionals who must be prepared to manage potential adverse reactions.

The integration of artificial intelligence (AI) in DR screening has shown great potential, with AI models capable of identifying disease patterns in retinal images, thus streamlining the diagnostic process and alleviating the burden on ophthalmologists (Malerbi et al. 2021, Sosale 2019). However, the quality of the images used to train these AI models remains critical to their accuracy (Nakayama et al. 2024). Notably, the presence or absence of mydriasis can significantly impact image clarity and, by extension, the performance of AI-based diagnostics with portable cameras.

This article investigates the impact of mydriasis on image gradability and explores factors associated with lower image quality, as well as its effect on the performance of AI models in DR screening. By comparing diagnostic accuracy between mydriatic and non-mydriatic images, this study aims to clarify how pupil dilation influences the reliability of AI systems for detecting retinal diseases, focusing on resource-limited settings where portable fundus cameras are becoming more widely used.

## Methods

This study included patients who underwent retinal exams both before and after mydriasis with Tropicamide 0.5% eyedrops; the imaging protocol consisted of two images captured per eye (macula-centered and optic disc-centered) by a variety of healthcare professionals, including ophthalmic technicians and certified ophthalmologists. Age, gender, self-reported race, and clinical comorbidities data were collected and included in this analysis.

Two masked, certified ophthalmologists independently conducted labeling. In cases of disagreement, a third senior retinal specialist provided adjudication. Diabetic retinal lesions—including hemorrhages, microaneurysms, venous beading, intraretinal microvascular abnormalities, new vessels, vitreous or preretinal hemorrhage, and retinal tractional membranes—were assessed based on the ICDR score (Wilkinson et al. 2003).

For assessing gradability, images were classified by the human labelers when at least 2/3 of the retina was visible, and image focus allowed them to assess the third branch of vessels as gradable (Nakayama et al. 2023).

### Device Description

The Phelcom Eyer (Phelcom Technologies, LLC, Boston, MA) is a portable retinal camera that integrates with a Samsung Galaxy S10 smartphone running on Android 11. The device is designed to capture high-resolution retinal images and facilitate diabetic retinopathy screening. It has a 12-megapixel sensor that produces images with a resolution of 1600 × 1600 pixels, ensuring detailed visualization of the retina. The camera’s 45° field of view allows for wide-angle fundus photography, which is essential for comprehensive retinal screening. Additionally, the Eyer features an autofocus range of −20 to +20 diopters, enabling precise focusing across different refractive states of the eye. The device may be used for handheld capture as well as slit lamp-based imaging.

### Artificial intelligence algorithm

We employed a convolutional neural network (CNN) pre-trained on ImageNet, for the classification of diabetic retinopathy. The model was trained on the mBRSET dataset (Nakayama et al. 2024) and validated on the study dataset.

Each image was labeled as either “normal” or “diabetic retinopathy” based on the International Clinical Diabetic Retinopathy (ICDR) scale (Wilkinson et al. 2003). Preprocessing steps included resizing the images to 224×224 pixels, normalization using standard ImageNet values, and random augmentations such as flips and rotations to increase the robustness of the model.

### mBRSET

The mobile Brazilian Ophthalmological dataset (mBRSET) conducted during the Itabuna Diabetes Campaign in Bahia, Brazil, involved 1,291 patients and collected 5,164 retinal fundus images, obtained after pharmacological mydriasis, using the Phelcom Eyer (Nakayama et al. 2024). The dataset includes patients with diverse backgrounds, reflecting Bahia’s mix of European, African, and Native American ancestry. The average patient age was 61.4 years (SD 11.6), and 65.1% were female, with a 23.2% rate of DR-positive exams (Wu et al. 2024).

### Model Training and Validation

The ResNet-200d architecture was fine-tuned for binary classification (normal vs. DR) using cross-entropy loss and optimized with the Adam optimizer. The dataset was split into an 80/20 ratio for training and testing. Training was conducted over 50 epochs with a batch size of 4.

Performance metrics such as accuracy, F1 score, area under the receiver operating characteristic curve (AUC), and confusion matrices were computed for each subgroup (mydriatic vs. non-mydriatic).

### Performance Evaluation and Subgroup Analysis

A univariate logistic regression was performed to assess the impact of demographics and clinical comorbidities on gradability. A Chi-square test was performed to assess the statistical significance of the differences in performance between the two groups, with a p-value threshold of <0.05 used to determine statistical significance.

To assess the performance of the AI algorithm in identifying positive DR cases, we analyzed pairs of gradable images under both mydriatic and non-mydriatic conditions. Model performance metrics, including accuracy, F1 score, AUC, and confusion matrix values (true positives [TP], false positives [FP], true negatives [TN], and false negatives [FN]), were calculated and compared across the mydriatic and non-mydriatic groups. Additionally, we conducted a focused analysis on performance discrepancies, specifically examining the rates of false positives and false negatives in each condition.

## Results

The study analyzed retinal fundus photographs from 327 patients, with an average age of 57.03 years (SD 16.82, ranging from 9 to 90 years), of whom 45.26% were male. Baseline demographics and comorbidities are presented in Table 1. At the patient level, 44% had no signs of retinopathy, 26.47% were diagnosed with non-proliferative diabetic retinopathy, and 29.31% had proliferative diabetic retinopathy.

**Table 1.**
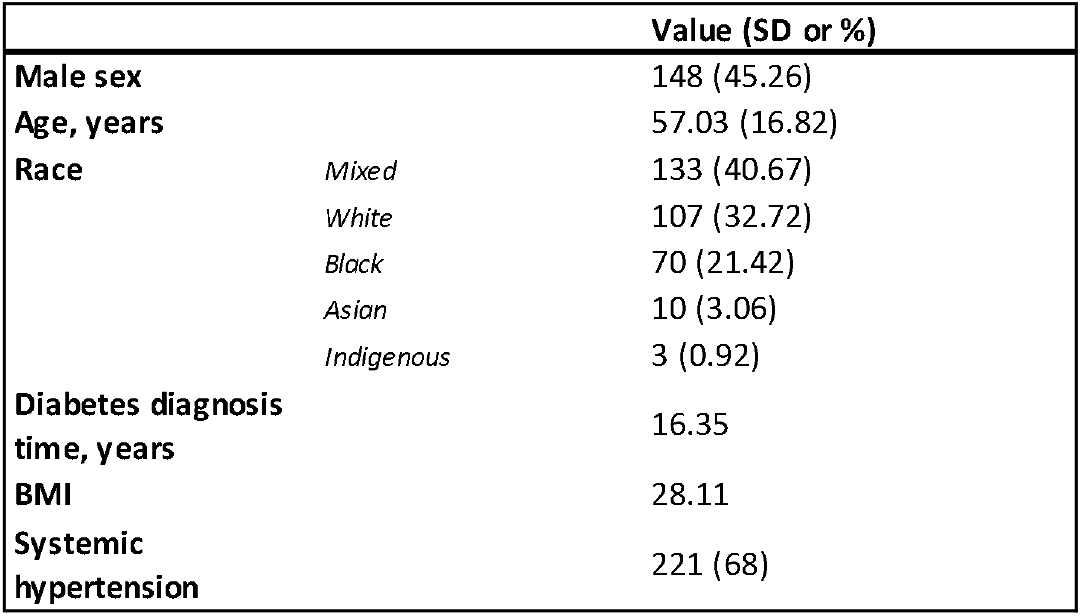
Description of demographics and the clinical comorbidities of study participants.

In the mydriasis group, 1056 images (82.1%) were deemed gradable, compared to 699 images (55.6%) in the non-mydriasis group, with a statistically significant difference (P < 0.001). (Figure 1)

**Figure 1:**
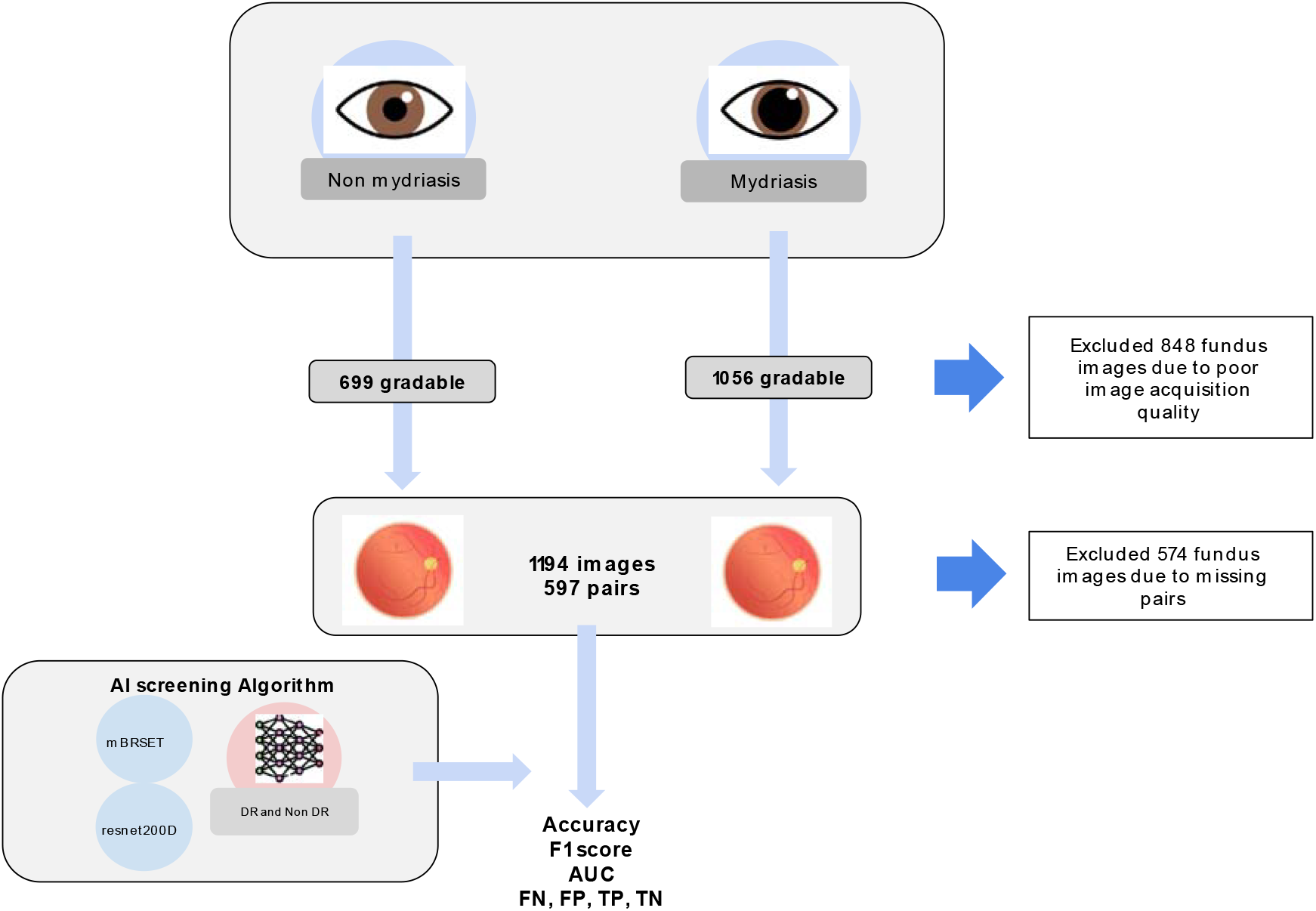
Flowchart of the study with the number of included and excluded images.

In the non-mydriasis group, systemic hypertension, age, duration of diabetes, and male gender were all significantly associated with a decreased likelihood of obtaining gradable retinal images. Systemic hypertension had the largest effect, with hypertensive patients showing a notable decrease in image quality (P < 0.0001). Age also showed a strong negative association, with each additional year slightly reducing the odds of high-quality images by 3.2% (P < 0.0001). Similarly, a longer duration of diabetes was linked to a reduced likelihood of high-quality images, with each additional year lowering odds by about 2.8% (P < 0.0001). Male gender was also a significant factor, associated with lower odds of high-quality images compared to females (P < 0.0001).

In terms of race, Indigenous patients were significantly associated with lower image quality (p = 0.049), suggesting potential disparities. In contrast, Black and Mixed-race categories did not show significant associations. (Table 2)

**Table 2.** Univariate logistic regression analysis for image quality.

| Variable | Coefficient | P-value |
| --- | --- | --- |
| Systemic Hypertension | -7.065 | <0.0001 |
| Age | -0.0322 | <0.0001 |
| Duration of Diabetes | -0.0276 | <0.0001 |
| Gender (Male) | -0.4647 | <0.0001 |
| Race (Black) | -0.1281 | 0.3568 |
| Race (Indigenous) | -1.3162 | 0.049 |
| Race (Mixed) | -0.0877 | 0.451 |
| Race (White) | 0.2207 | 0.072 |

### Artificial intelligence

The classification models showed a baseline performance of 83.03% accuracy, an F1 score of 0.83, and an AUC of 0.93 in the mBRSET testing subset, using a threshold of 0.5.

In the study dataset, after the image quality screening, 1194 images were included, consisting of 597 pairs of non-mydriatic and mydriatic images. In the mydriatic group, the model achieved an accuracy of 82.91%, compared to 79.23% in the non-mydriatic group for detecting cases of DR. The F1 score and AUC were higher in the mydriatic group, with values of 0.83 for F1 and 0.94 for AUC, compared to 0.79 and 0.93 in the non-mydriatic group.

The mydriatic group had fewer false positives (83 FP, 13.9%) compared to the non-mydriatic group (106 FP, 17.8%), the number of false negatives was higher in the mydriatic group (19 FN, 3.2%) compared to the non-mydriatic group (18 FN, 3.0%) (Figure 2). The lower false positive rate in the mydriatic group indicates that the model was less likely to incorrectly identify disease in patients without DR when mydriasis was applied.

**Figure 2:**
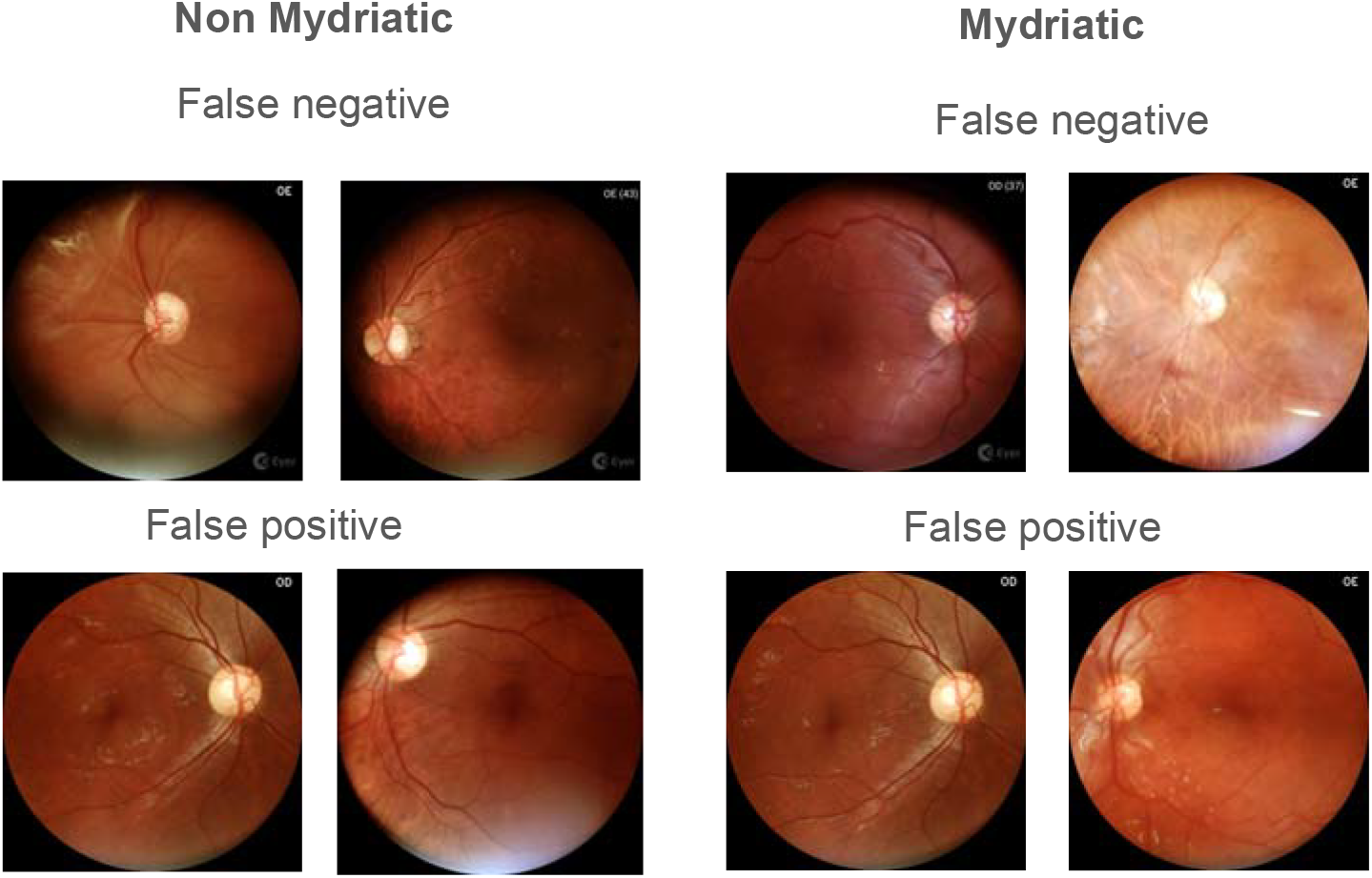
Examples of non-mydriatic and mydriatic false positives and false negatives exams.

The Chi-square test results showed no statistically significant difference between the mydriatic and non-mydriatic groups (P= 0.3279).

## Discussion

Diabetic retinopathy is a global challenge, with rising numbers, particularly affecting LMICs (Sivaprasad & Pearce 2019). For diabetic retinopathy screening programs, using non-mydriatic photos can be more pragmatic, with comparable results in detecting referable cases (Piyasena et al. 2018). This approach is particularly beneficial in resource-limited settings where healthcare professionals are not always available to administer mydriatic eyedrops and where managing rare but potential adverse effects of these agents can pose additional challenges (Sosale 2019). Previous studies have reported 19.7% (Scanlon et al. 2003) and 26% (Murgatroyd et al. 2004) of ungradable images with tabletop cameras. However, our study found a higher rate of ungradable images—44.4%—using a portable camera in non-mydriatic images.

Consistent with previous findings, our study shows that older age is associated with poorer image quality (Scanlon et al. 2005). Notably, we also report correlations with male gender, systemic hypertension, and the duration of diabetes mellitus diagnosis. While portable cameras are becoming increasingly popular in LMIC due to their being more affordable and technically easier to use (Rajalakshmi et al. 2021, Wu et al. 2024), they pose greater challenges in terms of image quality and gradability compared to tabletop cameras.

The use of automatic DR screening is promising for limited resource settings; nevertheless, image quality is an important factor that impacts the performance of automated models when deployed to clinical practice. Nevertheless, non-mydriatic images from portable cameras lead to more artifacts and worse image quality, with possible downstream consequences when employing automated screening.

The findings of this study emphasize the impact of mydriasis on retinal imaging for AI-based DR screening. In the mydriatic group, the model demonstrated improved accuracy F1 score, and AUC compared to the non-mydriatic group, indicating that pupil dilation may enhance model performance in DR detection. Additionally, the mydriatic group had fewer false positives compared to the non-mydriatic group, suggesting a reduced likelihood of incorrectly identifying DR in patients without the disease when mydriasis was applied.

These results underscore the potential benefit of mydriasis in improving image quality and diagnostic precision, especially in settings where high-quality images are crucial for early DR detection. The lower false positive rate in the mydriatic group suggests that pupil dilation may help reduce overdiagnosis, enhancing the model’s specificity.

In LMICs, where access to follow-up care may be limited, the lower number of false positives with mydriasis could help reduce unnecessary referrals, making AI-based screening more efficient. Balancing reduced false positives with a low rate of false negatives is essential to maximize the effectiveness of AI in DR screening for these settings. Moreover, the variability in the availability of mydriatic agents in LMICs presents another challenge. In some regions, mydriasis may not be feasible due to cost, patient discomfort, or lack of access to the necessary pharmacological agents. In such cases, non-mydriatic imaging may be the only option, despite its potential to produce lower-quality images. This variability in imaging conditions could affect the consistency of screening results and complicate the implementation of AI models in real-world settings.

This study has several limitations. First, there is a learning curve associated with acquiring images using handheld portable cameras. Although the healthcare professionals involved had experience with the camera, they were not specialized in retinal imaging, which could affect image quality and consistency. Additionally, by focusing solely on pairs of images, there is a selection bias, as this approach may not capture the full variability of image quality across all non-mydriatic images. This reduced sample size for non-mydriatic images may impact the comparison between mydriatic and non-mydriatic groups, possibly skewing performance assessments.

Furthermore, the generalizability of our findings is limited by the use of a single dataset and a single camera model, which may restrict the applicability of the results to other clinical settings, particularly those employing different cameras or imaging protocols. Lastly, the analysis of combined referral groups misses the granularity of the DR classification prediction score accuracy. Future studies should aim to validate these findings across a wider range of datasets and camera models to ensure the broader applicability of AI-based diabetic retinopathy screening solutions.

The study also underscores the need for further research into optimizing AI models for different imaging conditions. While mydriasis improves overall performance, efforts should be made to reduce the rate of false positives and negatives, particularly in resource-limited environments where access to ophthalmological care is already constrained. Future studies should explore strategies for enhancing model performance in non-mydriatic images, potentially through advanced image preprocessing techniques or developing specialized algorithms for low-quality images.

## Data Availability

The scripts underlying this study can be found on our GitHub repository: https://github.com/luisnakayama/portable_mydriasis.

